# Immunological pathways underlying autism: Findings from Mendelian randomization and genetic colocalisation analyses

**DOI:** 10.1101/2022.02.16.22271031

**Authors:** Christina Dardani, Jamie W. Robinson, Jie Zheng, Aws Sadik, Panagiota Pagoni, Evie Stergiakouli, Renee Gardner, Alexandra Havdahl, Jakob Grove, the iPSYCH Autism Spectrum Disorder working group, George Davey Smith, Sarah Sullivan, Beate Leppert, Hannah J. Jones, Stan Zammit, Golam M. Khandaker, Dheeraj Rai

## Abstract

Emerging evidence implicates the role of inflammation and immunity in autism. However, little is known about the involvement of specific immunological pathways and their causal role. In 18,381 autism cases and 27,969 controls from the PGC and the iPSYCH consortia, we investigated whether 15 cytokines implicated in the differentiation and function of CD4^+^ T cell subsets (T_H_1, T_H_2, T_H_9, T_FH_, T_H_17, T_Reg_) could be causally linked to autism. Within a Mendelian randomization framework, we used protein quantitative trait loci (pQTLs; N=1,000-3,394) to assess the effects of genetically proxied levels of plasma cytokines on autism. We additionally used brain cortex expression quantitative trait loci (eQTLs; N= 6,601) to investigate whether genetically predicted expression of the genes encoding the cytokines of interest influence autism. We performed colocalisation to assess the possibility that the identified effects were confounded due to Linkage Disequilibrium (LD). We also assessed the possibility of reverse causation. We report consistent evidence for causal effects of genetically predicted levels of IFN-γR1, IL-12Rβ1 (T_H_1), and IL-4RA, IL-5RA, IL-13RA1 (T_H_2) on autism. We identified brain-specific effects of genetically predicted expression of *IFNGR1, IL12RB1, IL23A*, which in the case of *IFNGR1* and *IL23A* were additionally supported by evidence suggestive of colocalisation. Findings appeared unlikely to be influenced by reverse causation. Our findings are consistent with a potentially causal effect of T_H_1 and T_H_2 pathway cytokines in autism, and further research is required to elucidate the pathways via which T_H_1 and T_H_2 influence its phenotypic presentation.

## INTRODUCTION

Autism is a common neurodevelopmental condition (∼1.5% worldwide prevalence^1^), influencing multiple areas of functioning across the life course^2^, and is associated with several comorbidities^3^, poor life outcomes^4^ and early mortality^5^. Little is currently known about the biological pathways contributing to autism and elucidating them may enhance current understanding on the phenotypic presentation, comorbidities, and life outcomes of autistic individuals.

Emerging evidence implicates inflammation and immunity in autism^6–8^. Nationwide registry-based studies suggest associations between parental autoimmune conditions, as well as maternal infections during pregnancy, and offspring autism^9–11^. Moreover, atypical levels of acute phase proteins (indicators of immune response^12^) in maternal serum during the first trimester of pregnancy, and in neonatal blood spots have been reported to be associated with autism^13,14^. Finally, two recent meta-analyses of case-control studies have provided evidence of atypical concentrations of pro- and anti-inflammatory cytokines in plasma and serum of autistic individuals versus controls^15,16^.

Cytokines are pleiotropic proteins^17^. Their key immune role includes driving differentiation of naïve CD4^+^ T cells into specific subsets, which are characterised by distinct cytokine products and functions^18,19^ (T helper 1 (T_H_1), T helper 2 (T_H_2), T helper 9 (T_H_9), T follicular helper (T_FH_), T helper 17 (T_H_17) and regulatory T cells (T_Reg_); please see Table 1 for an overview of inductive and product cytokines as well as subset functions). CD4^+^ T cells are types of T lymphocytes and are orchestrators of anti-viral, autoimmune, and anti-tumour responses^18–21^.

**Table 1.** Summary of the six major CD4^+^ T cells, the cytokines inducing their differentiation, their product cytokines and each subset functions*.

| CD4<sup>+</sup> T cell subsets | T<sub>H</sub>1 | T<sub>H</sub>2 | T<sub>H</sub>9 | T<sub>FH</sub> | T<sub>H</sub>17 | T<sub>Reg</sub> |
| --- | --- | --- | --- | --- | --- | --- |
| Inductive Cytokines | IL-2<br>IL-12 | IL-2<br>IL-4 | IL-2<br>IL-4<br>TGFβ | IL-6<br>IL-21 | IL-6<br>IL-21<br>IL-23<br>TGFβ | IL-2<br>TGFβ |
| Product Cytokines | IFN-γ | IL-4<br>IL-5<br>IL-13 | IL-9 | IL-21 | IL-17A<br>IL-17F<br>IL-22 | IL-10<br>TGFβ |
| Subset functions* | Macrophage activation<br>Inflammatory response against intracellular pathogens | Eosinophil activation<br>Allergic and autoimmune response | Response in helminth infections<br>Allergic and autoimmune response<br>Anti-tumour immune response | B cell activation<br>Inflammatory response against extracellular pathogens | Neutrophil activation<br>Inflammatory response against extracellular pathogens<br>Autoimmune response | Regulation of inflammatory response<br>Regulation of autoimmune response<br>Suppression of anti-tumour immune response |

\*The table summarises some of the main functions of the CD4<sup>+</sup> T cell subsets and does not imply that these are the only immune pathways and processes that they have been found to be implicated. A detailed description of each subset, signature cytokines and functions can be found in relevant publications<sup>18,19,75–78</sup>. IFN-γ: Interferon gamma; IL2-23: Interleukins 2-23; TGFβ: Transforming growth factor beta; T<sub>H</sub>1: T helper 1; T<sub>H</sub>2: T helper 2; T<sub>H</sub>9: T helper 9; T<sub>FH</sub>: T follicular helper; T<sub>H</sub>17: T helper 17; T<sub>Reg</sub>: Regulatory T cells.

There is increasing evidence suggesting the involvement of CD4^+^ T cell subsets and their signature cytokines in pregnancy outcomes, including recurrent miscarriages and preeclampsia^22^. In the case of autism, rodent studies suggest an association between maternal T_H_17 cell populations and their product cytokine IL-17A, and offspring autism-like behaviours^23^. Furthermore, observational studies have found atypical levels of T_H_1 and T_H_2 cytokines in neonatal blood spots^24^ as well as brain tissue^25^ of autistic individuals, while there are indications of associations between T_H_17, T_H_2 and T_Reg_ cell populations and gastrointestinal symptoms in autism^26^. Despite evidence suggesting a potential link between CD4^+^ T cells, their signature cytokines, and autism, the question of causality has yet to be settled, as residual confounding and reverse causation remain key challenges of current observational evidence.

In the present study we investigated the causal influence of genetically proxied cytokines implicated in the differentiation and function of six major CD4^+^ T cell subsets (T_H_1, T_H_2, T_H_9, T_FH_, T_H_17, T_Reg_) on autism, to elucidate potential distinct immunological mechanisms underlying the condition. We implemented Mendelian randomization (MR), through an instrumental variables approach, using single nucleotide polymorphisms (SNPs) associated with plasma cytokines (protein quantitative trait loci-pQTLs)^27–30^ as instruments. The method, under certain assumptions, intends to estimate causal effects and can overcome some of the limitations of observational studies, particularly reverse causation and residual confounding^31^. To gain insights into potential brain-specific effects, we additionally performed MR using SNPs associated with the expression of genes encoding the cytokines of interest in the brain cortex (expression quantitative trait loci-eQTLs)^32^. We complemented MR findings by performing genetic colocalisation analyses to provide evidence for the presence of shared causal variant(s) influencing levels/expression of the exposure (cytokine) and autism risk, meaning there might be a shared underlying biological mechanism^33,34^. Finally, we assessed the possibility of bias due to reverse causation by performing Steiger filtering^35^ and bidirectional MR analyses (genetic liability to autism influencing circulating cytokines)^36^.

## MATERIALS AND METHODS

A summary of the analysis plan can be found in Figure 1.

**Figure 1.**
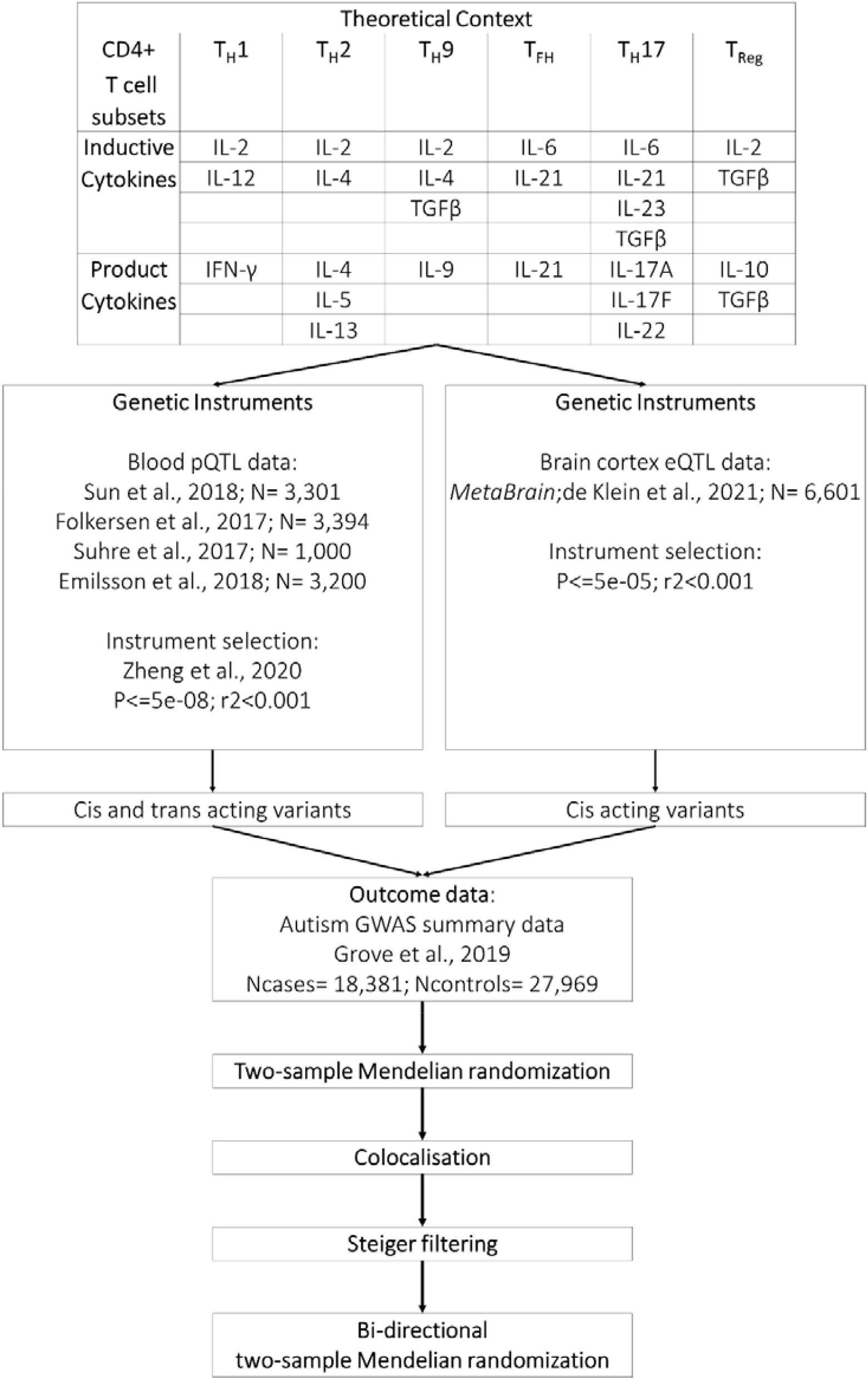
Summary of the analysis plan followed in the present study.

### Data sources and instrument definition

#### Blood plasma pQTL data

Plasma pQTL data for 15 of the cytokines of interest (Table1) were available in four genome-wide association studies (GWAS): Sun et al, 2018 (N= 3,301)^27^, Folkersen et al., 2017 (N= 3,394)^29^, Suhre et al., 2017 (N= 1,000)^28^, Emilsson et al., 2018 (N= 5,457)^30^. Supplementary Table S1 summarises sample sizes, population ancestry and methods for phenotype definition for each study. Further details on participants, plasma protein measurements, and genotyping of each study, can be found in the original publications.

In previous published work by Zheng et al^37^, genetic instruments across these GWASs were validated in terms of their consistency (limited agreement of pQTL association estimates across studies might indicate artefactual associations and therefore violation of the first MR assumption-Figure 2) and their specificity (pQTLs associated with several proteins can indicate pleiotropy and therefore potential violation of the third MR assumption-Figure 2). Further details on the validation protocol can be found in the original publication^37^. We considered this information important for the appraisal of the MR findings and therefore extracted 18 pQTLs that were independent (r^2^<0.001; 10,000Kb) and robustly associated (p≤5*10^−08^) with the cytokines of interest and their receptors (Supplementary Table S2 for a summary of the pQTLs, effect sizes, standard errors, p-values, GWAS source, specificity and consistency across studies). The only exceptions were instruments for IL-4 Receptor Subunit Alpha (IL-4RA) and IL-17F which were not included in the Zheng et al.^37^ validation process and therefore were extracted (r^2^<0.001; 10,000kb; p≤5*10^−08^) from the Sun et al GWAS data^27^ along with information on their respective regions, necessary for subsequent colocalisation analyses.

**Figure 2.**
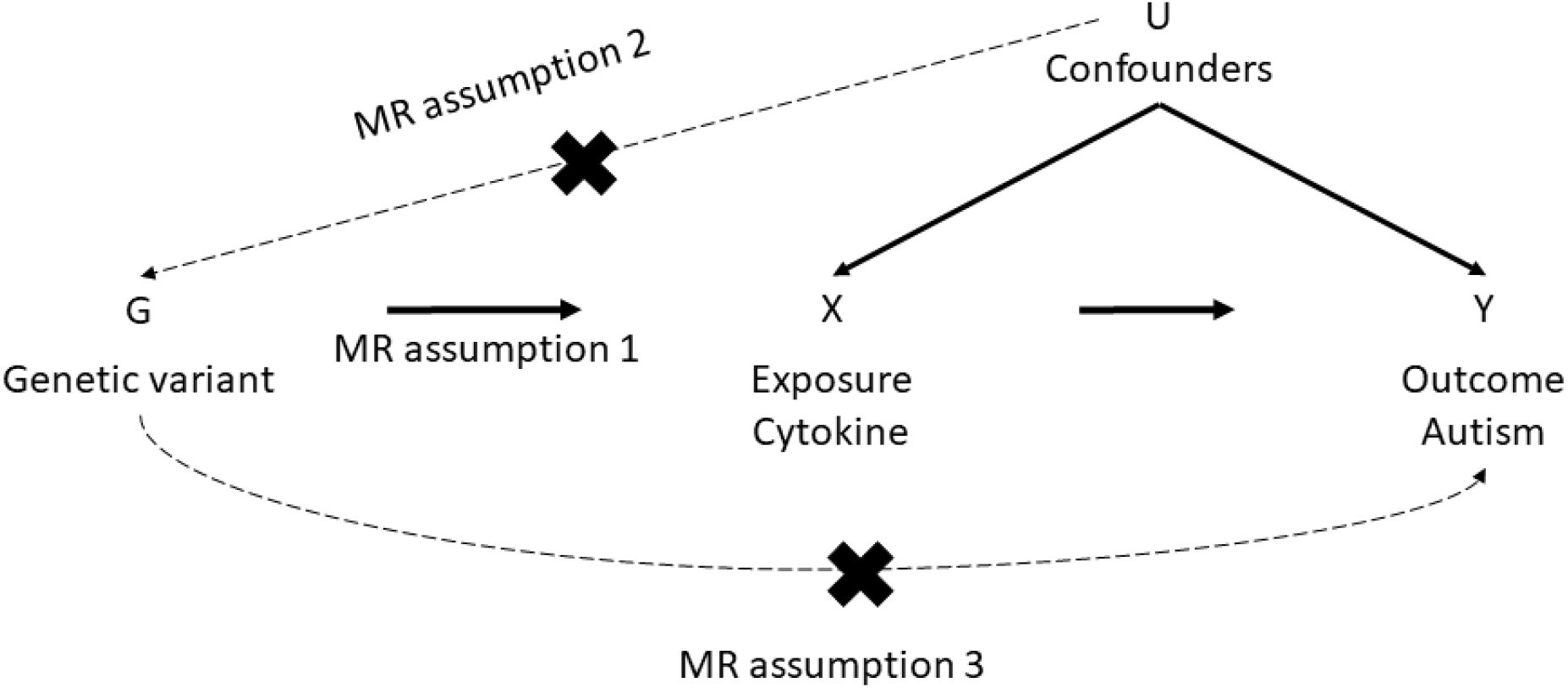
Directed acyclic graph (DAG) visualising the three MR assumptions. Specifically, the method can yield unbiased causal effect estimates under assumptions that the instruments should satisfy: they must be robustly associated with the exposure (MR assumption 1), they must not be associated with any confounders of the exposure-outcome associations (MR assumption 2), they should operate on the outcome entirely through the exposure (i.e. no horizontal pleiotropy).

All instruments were categorised into cis-acting and trans-acting (Supplementary Table S2). Instruments were categorised as cis-acting when they were located within proximity (±1Mb) to the cytokine-encoding gene, whereas instruments were categorised as trans-acting when located outside this window. SNPs acting in cis to the cytokine-encoding gene are more likely to influence mRNA and protein expression (thus being less pleiotropic)^38^. On the other hand, trans-acting SNPs, are more likely to be pleiotropic due to their distance from the cytokine-encoding gene, but their inclusion can potentially increase the proportion of variance explained in the exposure and the power of the MR analyses^37,38^.

#### Brain cortex eQTL data

Brain cortex eQTL data for the genes encoding the cytokines of interest were available in the largest meta-analysis of brain-derived eQTL datasets (*MetaBrain*), resulting in 6,601 RNA-seq samples Supplementary Table S1)^32^. Further details on the study datasets, samples and genotyping can be found in the original publication.

Cis-acting only eQTLs (±1Mb within the cytokine encoding gene region) were used for these analyses. This was because the *MetaBrain* study reported only the statistically significant trans-eQTLs, without information on the respective regions around them. This means that any genes with trans-acting SNPs are ineligible for subsequent colocalisation analyses. We defined as instruments SNPs that were independent (r^2^<0.001; 10,000 kb) and met a p-value threshold of 5*10^−08^. In cases that there were no instruments available for a cytokine of interest at this threshold, we used a relaxed p-value threshold of 5*10^−05^ in order to ensure that there would be at least one cis-acting eQTL for all the cytokine-encoding genes. In total, 19 eQTLs were extracted and details can be found in Supplementary Table S3.

#### Autism GWAS data

We used summary-level data from the latest autism GWAS of 18,381 cases and 27,969 controls^39^ (Supplementary Table S1). Considering emerging evidence suggesting that autism with co-occurring intellectual disabilities is distinct to autism without (in terms of behavioural characteristics^40^, genetic and environmental risk factors^41,42^, and comorbid medical and mental health conditions^43,44^), we additionally used summary-level data on a sub-sample of the iPSYCH cohort^45^ excluding all intellectual disability cases (cases= 11,203; controls= 22,555).

### Methods

#### Two-sample Mendelian randomization

MR utilises the special properties of germline genetic variants to strengthen causal inference within observational data^46^. Here we implemented this as an instrumental variables analysis using SNPs as instruments. The method can yield unbiased causal effect estimates under assumptions that the instruments should satisfy: (1) they must be robustly associated with the exposure, (2) they must not be associated with any confounders of the exposure-outcome associations, (3) they should operate on the outcome entirely through the exposure (i.e., no horizontal pleiotropy)^47^ (Figure 2).

For the present study, we performed two-sample MR, in which instrument-exposure and instrument-outcome effect sizes and standard errors were extracted from separate GWASs conducted in independent samples but representative of the same underlying population^48^.

We assessed the strength of each instrument by estimating their F-statistic, where an F statistic of >10 is indicative of adequate instrument strength^49^. SNP-exposure effect sizes and standard errors were extracted from the autism GWAS^39^, and their alleles were harmonised to ensure SNP-exposure and SNP-outcome effect sizes correspond to the same allele. The Wald ratio was used to generate causal effect estimates and the two term Taylor expansion to approximate standard errors, as all exposures were instrumented by a single SNP^32,50^. The same process was followed using as an outcome the iPSYCH autism sub-sample excluding all intellectual disability cases. Supplementary Tables S4, S5, S6, S7 contain information on the harmonised datasets used for each MR analysis. We did not apply correction for multiple testing because the cytokines assessed are organised into interacting subsets (Table 1), limiting therefore the definition of the number of independent tests. Instead, we orient the readers to appraise the study results in the context of their consistency across analyses^51^.

#### Genetic colocalisation

Colocalisation approaches can complement MR approaches by elucidating a distinct aspect of the identified causal relationship between an exposure and an outcome^34^. Specifically, colocalisation allows the assessment of the hypothesis that any identified causal effects are driven by the same causal variant influencing both exposure and outcome, instead of distinct causal variants that are in linkage disequilibrium (LD) with each other^33^. In practice, the approach is harnessing SNP coverage within the same specified locus for two traits of interest and tests whether the association signals for each trait at the specified locus are suggestive of a shared causal variant^33^.

For each MR result providing evidence of a causal effect, we tested for colocalisation between the genetically proxied exposure and autism. We extracted regions of SNPs within ±500KB around the instrumented SNP and implemented the algorithm described by Zheng et al^37^ to perform pairwise conditional and colocalisation (PWCoCo) analyses, which assesses all conditionally independent signals in the exposure dataset region against all conditionally independent signals in the outcome data. Genotype data from mothers in the Avon Longitudinal Study of Parents and Children (ALSPAC)^52,53^ cohort were used as the LD reference panel (N= 7,921; details on the ALSPAC cohort and available genotype data can be found in Supplementary Note S1). We ran these analyses using the default settings, as suggested by the authors in the original publications^33,37^. Evidence of colocalisation was considered an H4 (probability of both traits having a shared causal variant) ≥ 0.8, as has been suggested by the original authors. However, we observed post-hoc that many results showed high evidence of H0-H2, which assumes no evidence of a causal variant in either trait possibly due to underpowered datasets. On this basis, we considered as evidence suggestive of colocalisation a weighted H4≥ 0.8^54^., calculated as H4/(H3+H4) (the probability of having a shared causal variant for both traits weighted to the probability of having non-shared causal variants for each trait). The weighted H4 may provide evidence of colocalization in underpowered datasets but is reliant on the assumption that such a variant exists.

As a post-hoc analysis to PWCoCo and the weighted H4 approach, we also performed LD check^37^. Although the method was initially proposed to approximate colocalisation in cases that sufficient SNP coverage in the region was not available^37^, in the context of the present study it was implemented because the current autism GWAS yielded association signals in only three loci^39^ (assuming that these three loci are unlikely to be the only causal loci for autism and that future, larger, GWASs will reveal more information on the genetic architecture of autism). We assessed the LD between the instrumented SNP and the top 30 SNPs associated with autism in the test region (r^2^>0.8 with any of the strongest 30 SNPs for autism in the region approximating colocalisation).

#### Examination of reverse causation: Steiger filtering

We performed Steiger filtering to assess whether causal effect estimates were influenced by reverse causation^35^. The method assesses whether the genetic variants proxying for the exposure explain more variance in the outcome, which, if this is the case, suggests that the primary phenotype influenced by the variant is the outcome rather than the exposure.

#### Examination of reverse causation: Bi-directional two-sample MR

We assessed the causal effects of genetic liability to autism on levels of plasma cytokines. Ten independent (r^2^<0.01; 10,000 kb) SNPs for autism were extracted from the autism GWAS^39^ using a relaxed p-value threshold of ≤5*10^−07^ to increase the number of instruments included in the analyses (p≤5*10^−08^, r^2^<0.01; 10,000 kb, yielded only 2 SNPs) (Supplementary Table S8 for details on the effect sizes, standard errors and p-values of the autism instruments). They were then extracted from the Sun et al^27^ GWAS summary data for each cytokine of interest and their alleles were harmonised (Supplementary Table S9 for harmonised datasets). The primary method of analysis was the Inverse Variance Weighted (IVW)^55^. The consistency of the IVW effect estimates was assessed using the MR Egger regression^55^, the Weighted Median^56^ and the Weighted Mode^57^ methods. Details on each method can be found in Supplementary Note S2. Bi-directional MR analyses could not be performed using the *MetaBrain* dataset due to not having full genome-wide data available.

### Software

Analyses were carried out using the computational facilities of the Advanced Computing Research Centre of the University of Bristol (http://www.bris.ac.uk/acrc/). Brain cortex cis-eQTLs were extracted using the Summary-data-based Mendelian Randomization (SMR) package version 1.03 (https://cnsgenomics.com/software/smr/). The TwoSampleMR R package was used to conduct two-sample MR analyses, Steiger filtering and to construct LD matrices for LD check analyses (https://github.com/MRCIEU/TwoSampleMR). The PWCoCo algorithm was implemented using the Pair-Wise Conditional analysis and Colocalisation analysis package version 0.3 (https://github.com/jwr-git/pwcoco).

## RESULTS

### Two-sample Mendelian randomization

#### Causal effects of genetically proxied plasma cytokines on autism

All instruments proxying plasma cytokines had good strength (F>10, Supplementary Table S2).

We found evidence of a causal effect of genetically proxied Interferon Gamma Receptor-1 (IFN-γR1: OR= 1.15; 95%Cis: 1.03-1.29; p= 0.02), Interleukin 13 Receptor Subunit Alpha-1 (IL-13RA1: OR= 1.16; 95%Cis: 1.00-1.34; p= 0.04), Interleukin 4 Receptor Subunit Alpha (IL4-RA: OR= 0.81; 95%Cis: 0.65-0.99; p= 0.04) and Interleukin 5 Receptor Subunit Alpha (IL-5RA: OR= 0.91; 95%CIs: 0.83-1.00; p=0.05). There was sub-threshold evidence to suggest a causal effect of Interleukin 2 (IL2: OR= 1.14; 95%CIs: 0.99-1.32; p=0.07) and Interleukin 12 Receptor Subunit Beta-1 (IL-12Rβ1: OR= 1.03; 95%CIs: 0.99-1.07; p= 0.12). Analyses using the autism sub-sample excluding all intellectual disability cases, yielded comparable effect estimates. Results are summarised in Supplementary Tables S10 & S11 and visualised in Figure 3.

**Figure 3.**
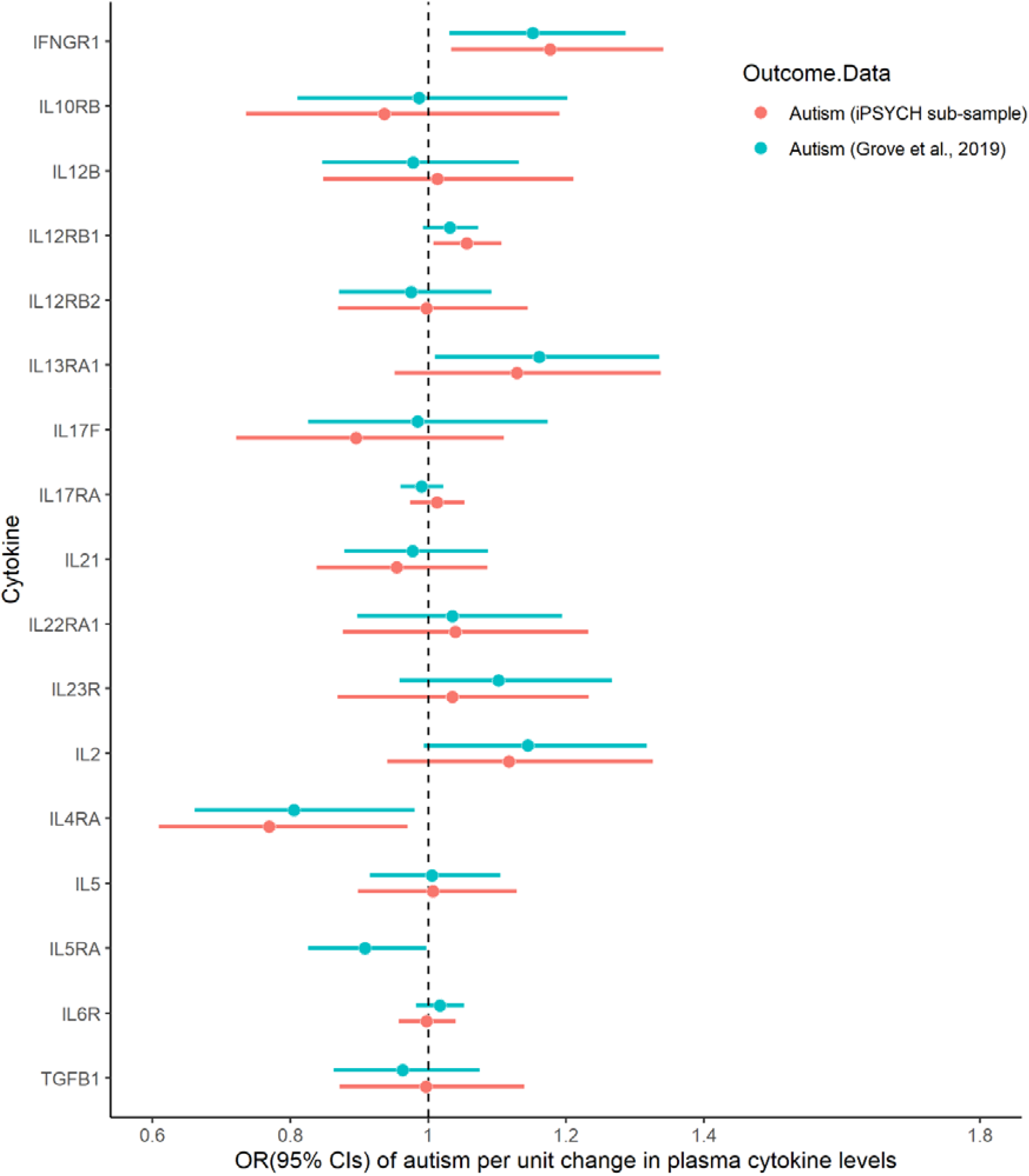
Forest plot of MR causal effect estimates and 95%CIs for autism per unit change in plasma cytokine levels. IFNGR1: Interferon Gamma Receptor-1; IL10RB: Interleukin 10 Receptor Subunit Beta; IL12B: Interleukin 12 Beta; IL12RB1: Interleukin 12 Receptor Subunit Beta-1; IL12RB2: Interleukin 12 Receptor Subunit Beta-2; IL13RA1: Interleukin 13 Receptor Subunit Alpha-1; IL17F: Interleukin 17 F; IL17RA: Interleukin 17 Receptor Alpha; IL21: Interleukin 21; IL22RA1: Interleukin 22 Receptor Subunit Alpha-1; IL23R: Interleukin 23 Receptor; IL2: Interleukin 2; IL4RA: Interleukin 4 receptor Subunit Alpha; IL5: Interleukin 5; IL6R: Interleukin 6 Receptor; TGFB1: Transforming Growth Factor Beta-1.

#### Causal effects of brain-expressed cytokine-encoding genes on autism

All instruments had good strength (F>10, Supplementary Table S3).

There was evidence to suggest a causal effect of genetically predicted expression of *IFNGR1* gene in brain cortex (OR= 1.22; 95%CIs: 1.05-1.42; p= 0.008), and *IL23A* gene (OR= 0.88; 95%CIs: 0.77-0.99; p= 0.04). There was suggestive evidence for a potential causal effect of genetically predicted expression of *IL12B* (OR= 1.24; 95%CIs: 0.97-1.57; p= 0.08) and *IL12RB1* gene (OR= 1.10; 95%CIs: 0.98-1.23; p= 1.11), which was stronger in subsequent analyses using the autism sub-sample excluding intellectual disability cases (*IL12B*: OR= 1.36; 95%CIs: 1.01-1.83; p= 0.04; *IL12RB1*: OR= 1.16; 95%CIs: 1.01-1.34; p=0.04). Results are summarised in Supplementary Tables S12 & S13 and visualised in Figure 4.

**Figure 4.**
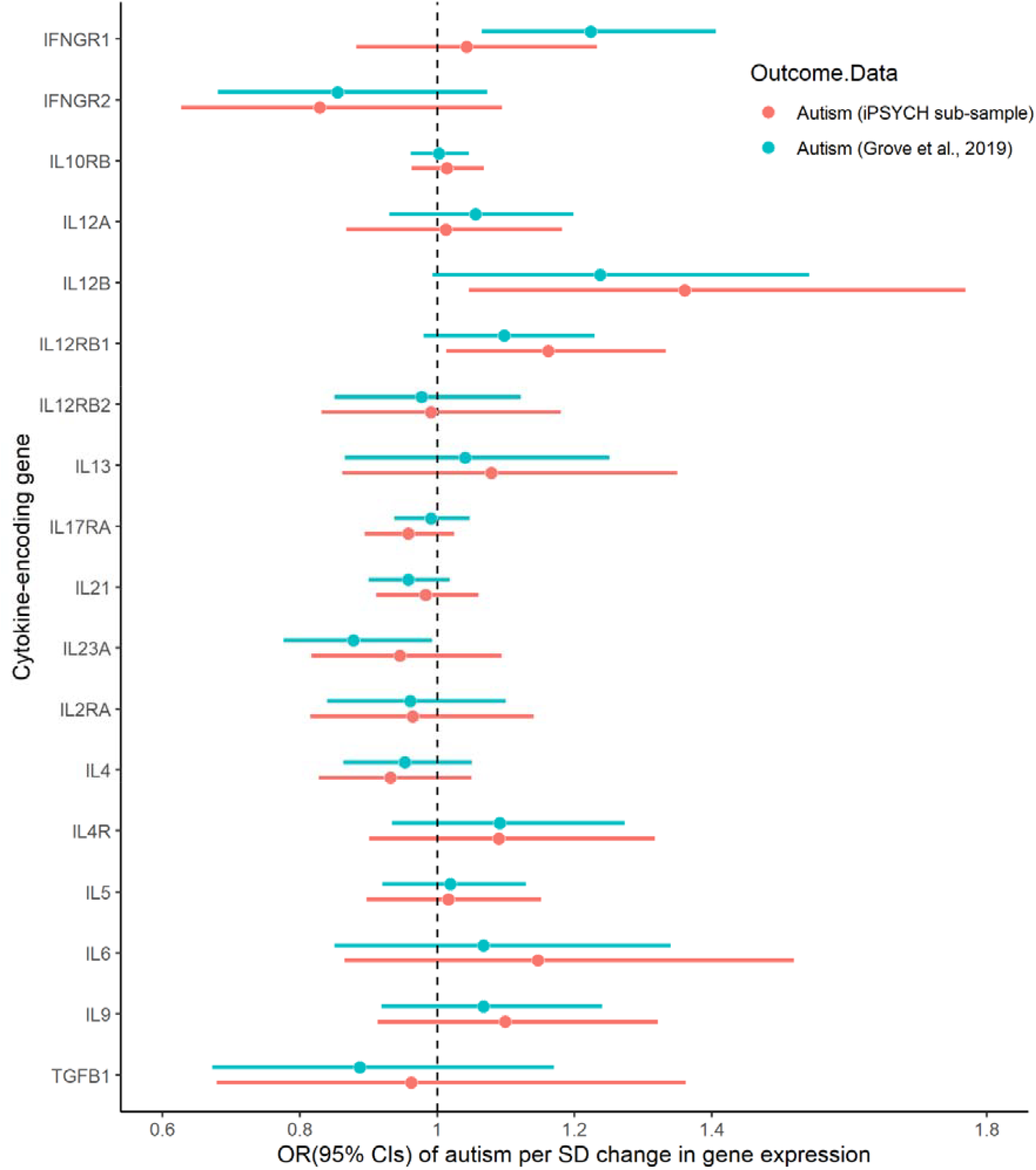
Forest plot of MR causal effect estimates and 95%CIs for autism per standard deviation change in cytokine-encoding gene expression in the brain cortex.

### Genetic colocalisation

None of the identified effects were supported by evidence of colocalisation i.e., H4≥0.8. However, there was evidence suggestive of colocalisation for the identified causal effect of genetically predicted expression of *IFNGR1* and *IL23A* in the brain cortex on autism. Specifically, the weighted H4 (H4/(H3+H4)) for both *IFNGR1* and autism as well as *IL23A* and autism was 0.9 and 0.8 respectively. In addition, LD Check analyses indicated that the lead *IFNGR1* and *IL23A* variants were in strong LD with at least one of the autism lead variants in the respective regions (r^2^>0.8) (Supplementary Table S14).

### Analyses to assess bias due to reverse causation

Steiger filtering indicated that across all analyses the genetic variants explained more variance in the exposure rather than the outcome, and that therefore the MR causal effect estimates were unlikely to be influenced by reverse causation (Supplementary Tables S10-S13).

We did not find any causal effect of genetic liability to autism on plasma cytokine levels (Supplementary Table S15).

## DISCUSSION

### Summary of findings

In the present study, we used MR and genetic colocalisation approaches to investigate the causal influence of genetically proxied cytokines implicated in the differentiation and function of six major CD4^+^ subsets (T_H_1, T_H_2, T_H_9, T_FH_, T_H_17, T_Reg_) on autism, and to elucidate potentially distinct immunological mechanisms underlying the condition. We found evidence suggesting a causal effect of genetically proxied T_H_1 (IFN-γR1, IL-12Rβ1), and T_H_2 (IL-4RA, IL-5RA, IL13-RA1) signature cytokines. There was additional evidence to suggest a causal effect of genetically predicted expression of *IFNGR1, IL12RB1* and *IL23A* genes in the brain cortex and especially in the case of *IFNGR1* and *IL23A*, the findings were supported by evidence suggestive of colocalisation. The identified effects appeared unlikely to be influenced by reverse causation bias.

### Findings in the context of existing evidence

There is a substantial body of evidence suggesting a potentially central role of T_H_1 and T_H_2 signature cytokines in autism. Specifically, in 1,100 neonatal dried blood spots from the Danish Newborn Screening Biobank, atypical levels of T_H_1 and T_H_2 cytokines (including IFN-γ, IL-2, IL-12, IL-4, IL-5) were found to be associated with later autism diagnosis^24^, while in 1,029 amniotic fluid samples from a Danish historic birth cohort, atypical levels of IL4 and IL5 were found to be associated with autism and broadly childhood neurodevelopmental and psychiatric disorders^58^. Although these findings could have been potentially influenced by selection bias (samples in both studies were restricted to pregnancies that amniocentesis was performed, i.e., high risk pregnancies), recent evidence from the population-based Early Markers for Autism (EMA) study further supports the potential role of T_H_1 and T_H_2 cytokines in autism as elevated concentrations of IFN-γ, IL-4 and IL-5 in maternal serum during gestation were found to be associated with offspring autism and intellectual disabilities^59,60^. A similar pattern has been identified in peripheral blood of children with autism, characterized by atypical levels of IFN-γ, IL2, IL4, IL5 and IL13^61,62^, as well as post-mortem brain tissue of adults with autism, characterised by increased levels of IFN-γ and an atypical IFN-γ/IL4 ratio^25^. By implementing the principles of MR and overcoming some of the limitations of previous studies (particularly reverse causation and residual confounding^31^), our study provides further support for a potentially causal role of genetically proxied T_H_1 and T_H_2 signature cytokines on autism. Especially in the case of IL-12Rβ1 and IL-5RA, we used cis-acting SNPs that were specific to the proteins, thus minimising the possibility of bias from pleiotropy. Similarly in the case of IFN-γR1, the instrument used showed specificity to the protein and consistency across pQTL studies.

### Potential immunological pathways in autism

Across MR analyses, a consistent pattern was identified for IFN-γR1. Elevated levels of genetically proxied IFN-γR1 as well as increased genetically predicted expression of *IFNGR1* gene in the brain cortex, were found to have causal links with autism. These findings were further supported by evidence suggestive of colocalisation, as well as evidence of causal effects of genetically predicted *IL12RB1* and *IL23A* in the brain cortex.

Specifically, IL-12Rβ1 (IL-12/23p40 subunit) is a common receptor for IL-12 and IL-23, which is promoting their signalling pathways^63^. However, the effect of IL-12Rβ1 on IL-12 and IL-23 signalling is not uniform. Increasing evidence suggests that IL12RB1 drives naïve CD4^+^ T cell differentiation to T_H_1 (IL12 pathway) or instead to T_H_17 (IL23 pathway), depending on the presence or absence of Interferon Regulatory Factor 1 (IRF1)^64^. IL12RB1, in the presence of IRF1, drives naïve CD 4^+^ T cell differentiation towards T_H_1 pathway leading to production of IFN-γ, whereas in the absence of IRF1, it drives differentiation towards T_H_17^65^. Our findings are consistent with this immunobiological understanding, as we identified evidence that autism is associated with increased genetically predicted expression of *IL12RB1* and *IFNGR1* in the brain cortex, but with decreased genetically predicted expression of *IL23A*.

Interestingly, the effects of *IFNGR1* and *IL23A* were pronounced in the autism sample including intellectual disability cases. IFN-γ signalling has been found to have a central role in brain function, influencing neurogenesis, synaptic plasticity and neurodegeneration^66^. Animal studies indicate that excess IFN-γ signalling and production drives neuronal cell death and synapse loss^67^, while epidemiological studies suggest associations between high circulating levels of IFN-γ and white matter damage in preterm infants^68^. This evidence might support our finding of a more pronounced effect of *IFNGR1* expression in brain cortex in autism cases including intellectual disabilities. However, given the sample sizes of the two autism GWASs we used, the possibility that our results reflect differences in power cannot be excluded.

### Strengths and limitations

The present study benefited from utilising a systematic approach for the selection of immune markers (based on CD4^+^ T cell subsets), as well as from the use of cis-acting genetic variants proxying for gene expression in the brain cortex. This allowed us to appraise our findings in the context of underlying immunological pathways and their mechanisms of action. Furthermore, we implemented a combination of MR and colocalisation approaches to strengthen causal inference and performed a series of sensitivity analyses to assess the possibility of reverse causation.

However, our findings should be appraised in the context of their limitations. First and foremost, none of the identified MR effects were supported by robust evidence of colocalisation. This might suggest that our MR findings were confounded due to LD. Although there was some evidence suggestive of colocalisation based on PWCoCo and post-hoc LD check analyses, this evidence relied on the assumption that there are causal variants in the regions of interest which is difficult to ascertain given the possibility of the datasets being underpowered. Future larger GWASs are necessary to further elucidate the present colocalisation findings. Second, some of the instruments used in our analyses were trans-acting, not specific to the cytokines of interest or consistent across studies and were selected using a relaxed p-value threshold. The inclusion of pleiotropic and weak instruments might have introduced bias in the causal effect estimates^69,70^. Third, although Steiger filtering suggested that our analyses were unlikely to be influenced by reverse causation, bi-directional MR analyses may have been underpowered considering the sample size of the outcome GWAS. Fourth, we assessed the contribution of common genetic variation and not rare, for which there is evidence of enrichment in immune-function gene sets in autism^71^. Fifth, we did not have access to family and individual level data which could have allowed the assessment of the origins of the identified effects (parental vs individual) as well as the possibility of non-linear effects (which can be particularly relevant in the case of immune response^72,73^). Sixth, autism is a highly heterogeneous condition and the possibility of distinct immunological pathways causally influencing autism subtypes was not possible to be assessed and cannot be excluded. Finally, analyses were conducted using summary data of European ancestry individuals, limiting therefore the generalisability of the present findings and replication across ancestries is necessary e.g., Zheng et al.,2021^74^.

### Conclusions

In conclusion, we found evidence consistent with a causal effect of genetically proxied T_H_1 and T_H_2 signature cytokines on autism. Particularly for IFN-γR1, there was additional MR and colocalisation evidence to suggest brain-specific effects of its respective gene expression on autism. The present findings appear unlikely to be influenced by reverse causation. Further research is necessary in order to elucidate the origins of the identified effects, the possibility of non-linear relationships and their possible distinct contributions across autism subtypes.

## Data Availability

Summary data used to conduct the present analyses can be found at: https://gwas.mrcieu.ac.uk/, https://metabrain.nl, https://www.med.unc.edu/pgc/download-results/. Summary data on autism excluding intellectual disabilities can be obtained after application to the iPSYCH Autism Spectrum Disorder working group.

## ETHICS DECLARATIONS

Across all analyses, summary-level data were used. Details on ethical approval and participant informed consent can be found in the original publications^27–30,32,39^. JWR receives funding from Biogen for research unrelated to the present study. No funding body has influenced data collection, analyses or their interpretation.

## ACKNOWLEDGEMENTS

The Medical Research Council (MRC) and the University of Bristol support the MRC Integrative Epidemiology Unit [, MC_UU_00011/1, MC_UU_00011/3, MC_UU_00011/5]. The UK Medical Research Council and Wellcome (Grant ref: 217065/Z/19/Z) and the University of Bristol provide core support for ALSPAC. GWAS data was generated by Sample Logistics and Genotyping Facilities at Wellcome Sanger Institute and LabCorp (Laboratory Corporation of America) using support from 23andMe. A comprehensive list of grants funding is available on the ALSPAC website (http://www.bristol.ac.uk/alspac/external/documents/grant-acknowledgements.pdf). This research was funded in part, by the Wellcome Trust. For the purpose of Open Access, the author has applied a CC BY public copyright licence to any Author Accepted Manuscript version arising from this submission. CD acknowledges the support of Wellcome Trust [215379/Z/19/Z]. GDS, HJ, DR, SS, SZ are supported by the NIHR Biomedical Research Centre at University Hospitals Bristol and Weston NHS Foundation Trust and the University of Bristol. The views expressed are those of the authors and not necessarily those of the NIHR or the Department of Health and Social Care. GMK acknowledges funding support from the Wellcome Trust (201486/Z/16/Z), the MQ: Transforming Mental Health (grant code: MQDS17/40), the Medical Research Council UK (grant code: MC_PC_17213 and grant code: MR/S037675/1), NIHR (project code: NIHR202646), and the BMA Foundation (J Moulton grant 2019). JWR is funded by Biogen. The iPSYCH team was supported by grants from the Lundbeck Foundation (R102-A9118, R155-2014-1724, and R248-2017-2003), NIMH (1U01MH109514-01) and the Universities and University Hospitals of Aarhus and Copenhagen. The Danish National Biobank resource was supported by the Novo Nordisk Foundation. High-performance computer capacity for handling and statistical analysis of iPSYCH data on the GenomeDK HPC facility was provided by the Center for Genomics and Personalized Medicine and the Centre for Integrative Sequencing, iSEQ, Aarhus University, Denmark. RG acknowledges funding support from the Swedish Research Council (VR2017-02900). AH was supported by grants from the South-Eastern Norway Regional Health Authority (2020022, 2018059) and the Research Council of Norway (274611, 288083).

